# Bridge Capture permits cost-efficient, rapid and sensitive molecular precision diagnostics

**DOI:** 10.1101/2024.04.12.24301526

**Authors:** Simona Adamusová, Anttoni Korkiakoski, Nea Laine, Anna Musku, Tuula Rantasalo, Jorma Kim, Juuso Blomster, Jukka Laine, Tatu Hirvonen, Juha-Pekka Pursiheimo, Manu Tamminen

## Abstract

Liquid biopsies are a less invasive alternative to tissue biopsies that have been the mainstay of cancer diagnostics to date. Recently, the quantification of mutations in circulating tumor DNA (ctDNA) by targeted next-generation sequencing (NGS) has been gaining popularity. Targeted NGS can be achieved through various library preparation methods, each with distinct advantages and limitations. Here we introduce Bridge Capture, a novel technology that goes beyond the advantages of market-leading liquid biopsy technologies, eliminating the need to compromise between scalability, cost-efficiency, sensitivity, or panel size. We compared Bridge Capture to leading commercial technologies currently available in cancer diagnostics; Archer™ LIQUIDPlex™ and AmpliSeq™ Cancer HotSpot Panel v2 for Illumina®. Of all methods, Bridge Capture detected the lowest mutant allele frequency (MAF) on matched contrived colorectal biospecimens mimicking ctDNA. Next, we demonstrated the capability of Bridge Capture to effectively utilize the sequencing capacity, permitting affordable and sensitive variant detection in smaller laboratories, while delivering significant cost savings for central laboratories. Additionally, we demonstrated the reproducibility of Bridge Capture by a high correlation between the results from two independent laboratories. These results highlight the method’s portability and suitability for kit use in entirely new settings. Moreover, we presented ease of automation and minimal hands-on time of Bridge Capture. Owing to its unique design, the Bridge Capture is compatible with the most commonly used NGS platforms. Taken together, Bridge Capture is cost efficient, simple, rapid and sensitive cancer diagnostics tool that significantly improves the detection of mutations in liquid biopsies.

## Introduction

Until recently, tissue biopsies have been the primary method for diagnosing and monitoring cancer. However, a new family of techniques, collectively known as liquid biopsies, is gaining popularity. Liquid biopsies, typically based on a simple blood draw, permit less invasive and simplified diagnostics, and more feasible longitudinal disease monitoring which is crucial for tracking the disease progression and the effectiveness of treatment over time (1). Moreover, liquid biopsies are unaffected by tumor heterogeneity – the spatial variation within a tumor – which is a downside for traditional biopsy (2).

A key component of liquid biopsies is circulating tumor DNA (ctDNA), which is a part of cell-free DNA (cfDNA) found in blood. ctDNA originates from tumor cells shedding their DNA either through apoptosis, necrosis, or active release and reflects the genetic makeup of the tumor in its entirety (2,3). A key aspect of ctDNA analysis is quantifying mutant allele frequencies (MAFs). A given MAF indicates the proportion of a mutation in cfDNA. A higher MAF generally suggests a larger tumor burden, providing insight into the extent of the cancer in the body (3–6).

Next-generation sequencing (NGS) is a crucial tool for precision cancer diagnostics and is roughly divided into whole genome sequencing (WGS) and targeted sequencing. WGS plays a pivotal role for reference building and variant discovery. When applied to cancer diagnostics, WGS is impractical due to high sequencing costs (typically 99.9 % of the reads obtained by WGS are irrelevant for cancer detection and prognosis) and complexity of data interpretation (7). Targeted sequencing, on the other hand, focuses only on the regions of interest relevant to the disease, therefore permitting more affordable sequencing costs through improved sequencing depth utilization, and decreased data storage and analysis requirements. Importantly, targeting of the sequencing effort permits detecting mutations present at very low MAFs (8).

Targeted sequencing can be achieved through various approaches such as amplicon-based, hybridization-based, and molecular inversion probe (MIP)-based NGS library preparation methods. Amplicon-based methods target regions of interest via specifically designed primer pairs by PCR (9,10). They provide a simple and rapid workflow (11–13), but generally have limited scalability due to primer cross-reactivity and are prone to false variant reads owing to increased error-rates coming from PCR amplification (7,14). Hybridization-based methods use chemically modified oligonucleotide probes to enrich pre-specified parts of NGS libraries (15–17). These methods permit extensive panels and can more efficiently provide information from difficult genomic regions such as repeat sequences (8). However, they are generally expensive and time-consuming. Molecular Inversion Probes (MIP) are single-stranded oligonucleotides that hybridize to target genomic region with terminal ends. The resulting gap between the probe ends is filled and the molecule is circularized (18). MIPs offer cost efficiency compared to hybridization-based methods, and improved panel size and scalability compared to amplicon-based methods. Their downsides include non-uniform coverage, high probe synthesis cost and increased noise compared to amplicon-based methods (19).

To address these shortcomings, we developed Bridge Capture, a novel targeted NGS library preparation technology that provides sensitive detection of low MAF while remaining rapid, highly multiplexed and cost-efficient. Bridge Capture uses oligonucleotide probe constructs to target specific genomic regions of interest (20) (**Fig. 1**). These constructs are composed of a bridge oligomer hybridized to left and right probes. After the construct hybridizes to the target of interest, all the gaps between the oligomers are filled and ligated, effectively capturing the target and creating a circular molecule. The bridge oligomer serves as a primer for initiation of Rolling Circle Amplification (RCA). The flowcell binding sequences are attached to the monomers through limited-cycle PCR. This unique design offers several benefits, namely allowing use of Bridge Capture with commonly available NGS platforms, decreasing the synthesis costs of the panels compared to MIP based methods, and permitting the use of highly multiplexed panels.

**Figure 1.**
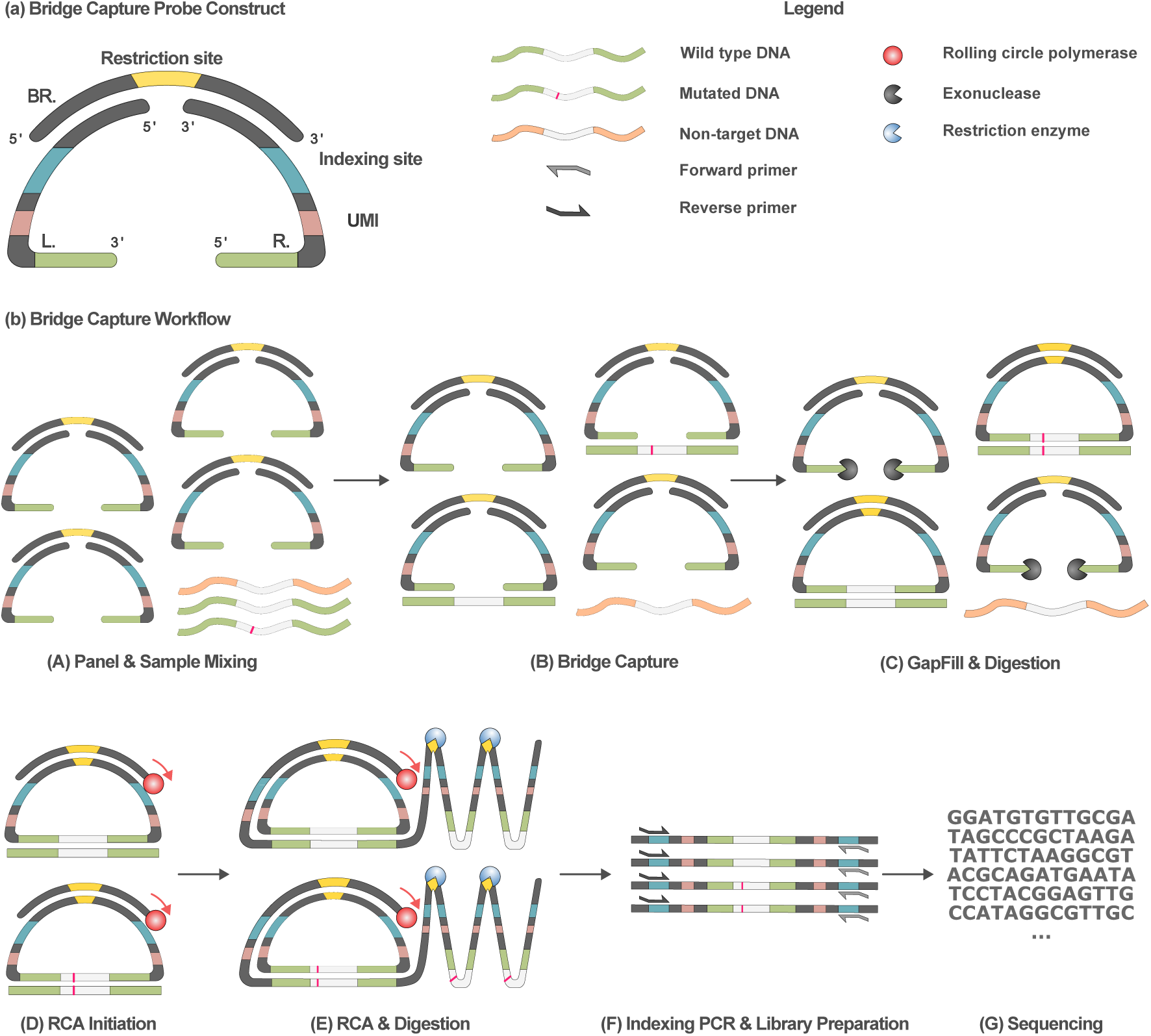
Bridge Capture in summary. (**a**) Bridge Capture probe construct consists of a bridge oligomer (BR.) with binding sites to left (L.) and right probe (R.). The bridge contains a restriction recognition site for a restriction enzyme (yellow). Left and right probe contain sequences for binding of dual index primers (blue), UMIs (pink) and target specific regions (green). (**b**) Bridge Capture workflow. The probe constructs are mixed with a sample containing sequences of interest (step A – Panel & Sample Mixing), to which the Bridge Capture probe constructs hybridize (step B – Bridge Capture). After Bridge Capture is complete, DNA polymerase and DNA ligase are used to fill all the gaps in the probe constructs to create circularized molecules. The constructs that failed to hybridize to target DNA are digested by addition of exonucleases (step C – GapFill & Digestion). RCA is initiated from the bridge oligomer of the circularized construct (step D – RCA Initiation). A long single stranded concatemer is synthetized by RCA and subsequently digested by a restriction enzyme creating multiple copies of a monomer (step E – RCA & Digestion). Newly produced monomers are indexed with dual indexing primers to create a library viable for sequencing (step F – Indexing PCR & Library Preparation). As the final part of the workflow, the library is sequenced (step G – Sequencing).

In this publication, we demonstrate analytical validation of Bridge Capture, a patented NGS library preparation workflow (20) that exceeds the performance of commercially available technologies while providing a workflow that combines the speed, sensitivity and simplicity of the amplicon- and MIP-based methods with the scalability of the hybrid capture methods.

## Materials and methods

### Sample preparation

Biospecimens were purchased from Indivumed GmbH (Hamburg, Germany), and consequently an ethics committee review was waived by the Research Ethics Committee of the wellbeing services county of Southwest Finland. Informed consent forms have been signed by each respective donor of the samples. Total DNA from fresh-frozen colorectal cancer (CRC) tissue samples was isolated using QIAamp DNA Mini Kit (50) (Qiagen, Hilden, Germany) and genomic DNA (gDNA) from the whole blood was extracted using NucleoSpin Blood (Macherey-Nagel, Düren, Germany). The extracted DNA was fragmented using Bioruptor® Pico (Daigenode, Liege, Belgium). 50 µL of sample was loaded in the 0.1 mL Bioruptor® Microtube (Diagenode) and sample was fragmented using 60 cycles with cycle conditions of 30”/30” to mimic size distribution of cfDNA. Processed samples were frozen at −80 °C and double-stranded DNA (dsDNA) concentration of the sample was measured the following day using Qubit 4 Fluorometer and Qubit dsDNA HS (high sensitivity) Assay Kit (ThermoFisher Scientific, Waltham, MA).

### Preparation of Bridge Capture probe panels

Bridge oligomer and probes were synthesized by Integrated DNA Technologies (IDT, Coralville, IA). Corresponding left and right probes were mixed with the bridge oligomer in 96-well plate using automatized pipetting robot Opentrons OT-2 (Opentrons, Brooklyn, NY) and afterwards annealed in C1000 Touch Thermal Cycler (Bio-Rad, Hercules, CA). Each annealed probe pair and bridge were pooled by Opentrons OT-2 to prepare 25 nM panel stock solution for 282-probe panel and 10 nM panel stock solution for 887-probe panel. The list of genes and the number of probes per gene in the 282-probe panel and in the 887-probe panel are provided in the **Supplementary Table 1** and **Supplementary Table 2**, respectively.

### Bridge Capture library preparation

To perform a Bridge Capture library preparation, a total of 320 ng of fragmented DNA (ca. 100,000 copies of gDNA) was used as starting material input. Samples underwent overnight (O/N) target capture using Genomill’s proprietary 282-probe panel (**Supplementary Table 1**). The details of Bridge Capture library preparation are further described in Pursiheimo et al. (20).The Bridge Capture libraries were purified with 0.9x Agencourt Ampure XP beads (Beckman Coulter, Brea, CA). The libraries were quantified by Qubit 4 Fluorometer and Qubit dsDNA HS (high sensitivity) Assay Kit (ThermoFisher Scientific). The sequencing platform utilized for each experiment is specified in its respective section.

### Technology comparison and inter-lab reproducibility

In order to compare Bridge Capture to other commercial technologies, two patient tissue CRC specimens were selected and underwent sample preparation. To confirm that the processed samples contained the six gene mutations, the samples were analyzed by Bridge Capture and were sequenced with iSeq 100 using iSeq 100 i1 Reagent v2 (300-cycle) 2 × 150 bp in paired-end run (Illumina). Presence of *APC* p.Q1406*, *KRAS* p.G13D, *PIK3CA* p.E545K and *APC* p.E1309Dfs*4, *KRAS* p.G12D, *TP53* p.Y126D was confirmed in sample 1 and sample 2, respectively. Patient samples were used as is or they were diluted with fragmented gDNA in various ratios to mimic different levels of MAFs in tenfold, hundredfold, and thousandfold dilution (**Supplementary Table 3**). Each sample was tested in three replicates for each technology tested: Bridge Capture, Archer™ LIQUIDPlex™ (from now on referred to as LIQUIDPlex) (IDT), and AmpliSeq™ Cancer HotSpot Panel v2 for Illumina® (from now on referred to as AmpliSeq) (Illumina). For Bridge Capture, 282-probe panel was used. The starting material load for Bridge Capture and LIQUIDPlex was 320 ng. To stay within the recommended load range, 100 ng of starting material was used for AmpliSeq. All the samples were processed by each technology at an independent diagnostic service provider. For LIQUIDPlex and AmpliSeq, sequencing was performed using Illumina NextSeq 500, and for Bridge Capture NextSeq 550 was used. NextSeq 500/550 High Output Kit v2.5 (300-cycle), 2 × 151 bp paired-end run (Illumina) was used for all three sequencings. Aliquots from the same samples were kept at Genomill for later processing by Bridge Capture to evaluate inter-lab reproducibility. Sequencing libraries prepared at Genomill were sequenced using the MiSeq - MiSeq Reagent Kit v3 for a 2 × 300 bp paired-end run (Illumina). Both laboratories processed the samples using a ready-made Bridge Capture kit, containing probe mixture, reagents, and protocol.

### Comparison of automated and manual Bridge Capture workflow

To evaluate the automated Bridge Capture workflow, two CRC specimens and fragmented gDNA were analyzed by automated and manually performed workflows. Detected SNVs included *KRAS* p.G12V, *APC* R876*, and *APC* E1408* in one CRC specimen, and *CTNNB1* p.S45P, *KRAS* p.G13D, *TP53* p.R248W in the other. Four sets of matched contrived samples were prepared in three replicates: undiluted CRC specimen 1 and 2, CRC specimen 1 and 2 diluted tenfold with fragmented gDNA, and fragmented gDNA (n = 15). One of the sets was analyzed following the manual Bridge Capture workflow protocol. The rest of the sets were analyzed in three separate runs by the automated workflow in 96-well plate using a pipetting robot Opentrons OT-2 and Opentrons Thermocycler GEN1 (Opentrons). For the manual workflow, all incubations were performed in C1000 Touch Thermal Cycler (Bio-Rad). The protocols used for the manual and the automated workflows were identical. The 320 ng of each sample was used and analyzed by 282-probe panel. The produced libraries were sequenced using NovaSeq 6000 SP2 2 × 150 bp flowcell (Illumina).

### Effect of the hybridization time on the Bridge Capture detection limit

To determine the effect of the hybridization time on the detection limit of Bridge Capture, 320 ng of diluted specimen was incubated with 282-probe panel under following incubation times: O/N, 4 hours, 2 hours, 1 hour and 0.5 hours. CRC specimen covered single-nucleotide variants (SNVs): *KRAS* p.G12V, *APC* R876* and *APC* E1408*. The specimen was diluted tenfold with fragmented gDNA, to simulate MAFs ten times lower than the original. The sample was tested in five replicates at each timepoint. Produced libraries were sequenced using the MiSeq system, with MiSeq Reagent Kit v3 for a 2 × 300 bp paired-end run (Illumina).

### Panel scalability

To evaluate the impact of increasing panel size on the evenness of the probe performance, the Bridge Capture analysis was performed with 282-probe panel and 887-probe panel. A total of 320 ng of fragmented gDNA was used as starting material. In the experiment, 10 technical replicates of fragmented gDNA were analyzed with both panels by Bridge Capture. Obtained libraries were normalized and sequenced using the MiSeq - MiSeq Reagent Kit v3 for a 2 × 300 bp paired-end run (Illumina).

### Data analysis and statistics

Bridge Capture reads were merged using VSEARCH (21) (v2.15.2_linux_x86_64) with the following parameters: --fastq_minovlen 10 --fastq_maxdiffs 15 --fastq_maxee 1 --fastq_allowmergestagger. A proprietary pipeline utilising Unique Molecular Identifier (UMI) based error-correction was used to process the Bridge Capture data. Both LIQUIDPlex and AmpliSeq data were processed by the independent diagnostic service provider using ArcherDX’s Archer Analysis Unlimited (v6.0.3.1) and Illumina’s DNA Amplicon (v2.1.1) data processing pipelines with default parameters (SNV cutoff LIQUIDPlex 0.5 %, Ampliseq 1 %). *In silico* subsampling was done without replacement to include 10 %, 1 % and 0.1 % of the original forward and reverse reads and then processed as mentioned before.

For the linear regression analysis of automated and manual workflow, three randomly selected replicas out of total nine replicas of automated workflow were paired with the three replicas of manual workflow.

For the figures and statistics, Python version 3.11.5 was used. The figures were drawn using matplotlib (v3.7.2) library, R^2^ scores were calculated using sklearn (v1.2.2) library, and scipy (v1.11.1) library was used for linear regressions, Spearman correlation and Analysis of variance (ANOVA). Statistical significance for panel performance was established by a permutation test (n=1,000,000) by calculating if the standard deviation of the ten replicas of a randomly selected probe in the panel was higher than the standard deviation of ten randomly selected signals (of any probe and of any replica).

## Results

### Bridge Capture

The hallmark of novel Bridge Capture method is the use of oligonucleotide probe constructs to target specific genomic regions of interest (20). The probe construct consists of a bridge oligomer annealed to left and right probes (**Fig. 1a**). The bridge includes sequences complementary to the probes, a restriction recognition site and various modifications protecting the bridge from exonuclease activity or unwanted elongation by polymerase. Left and right probes contain bridge binding sites, binding sites for sequencing platform specific adapters, UMI sequences, and most importantly the target specific binding sites.

In the first step of Bridge Capture, probe constructs are introduced to a sample containing the targets of interest (cfDNA/DNA isolated from blood or other sources; step A in **Fig. 1b**). Successful targeting by Bridge Capture construct results in a gap between the left and right target specific probes with the mutation of interest located within the gap region of targeted DNA strand (step B in **Fig. 1b**). Next, the gap is filled by DNA polymerase, starting from the 3’ end of left probe and continuing towards the 5’ end of right probe. The newly synthesized sequence is ligated to the 5’ end of the right probe by a DNA ligase. The gap between 5’ end of the left and 3’ end of the right probe, held together by a bridge oligo, is also filled by a DNA polymerase and ligated by a DNA ligase. This circularizes the probe construct and captures the mutation(s) of interest. Afterwards, all the non-circularized constructs are digested by exonucleases (step C in **Fig. 1b**). Rolling circle amplification is initiated from the 3’ end of bridge oligomer of circularized construct (step D in **Fig. 1b**). RCA generates multiple copies of the circular construct by creating a long concatemeric single-stranded DNA (ssDNA). After RCA, ssDNA concatemer is digested into monomers using a restriction enzyme (the restriction recognition site being provided in the bridge oligomer; step E in **Fig. 1b**). Sequencing platform specific adapters are introduced to the monomers through limited-cycle indexing PCR (step F in **Fig. 1b**). The resulting libraries are purified, pooled, and sequenced on the NGS platform of choice (step G in **Fig. 1b**). As of now, the Bridge Capture has been tested on several NGS platforms including Illumina NovaSeq, Ion Torrent S5 and Element Biosciences AVITI^TM^.

### Technology concordance

The performance of Bridge Capture was assessed by comparing it to commercially available cancer diagnostics technologies: LIQUIDPlex and AmpliSeq. The comparison was performed using dilutions of CRC biospecimens into fragmented gDNA (respective to genomic copies) to mimic different levels of MAFs, resulting in 24 samples in total (**Supplementary Table 3**).

The MAFs detected by the Bridge Capture were strongly correlated to the MAFs determined by commercially available technologies, with R^2^ = 0.995 between Bridge Capture and LIQUIDPlex, and R^2^ = 0.988 between Bridge Capture and AmpliSeq (**Fig. 2**). LIQUIDPlex did not target any *APC* mutations and AmpliSeq did not target *APC* p.Q1406*, and therefore these mutations were not included in the comparison.

**Figure 2.**
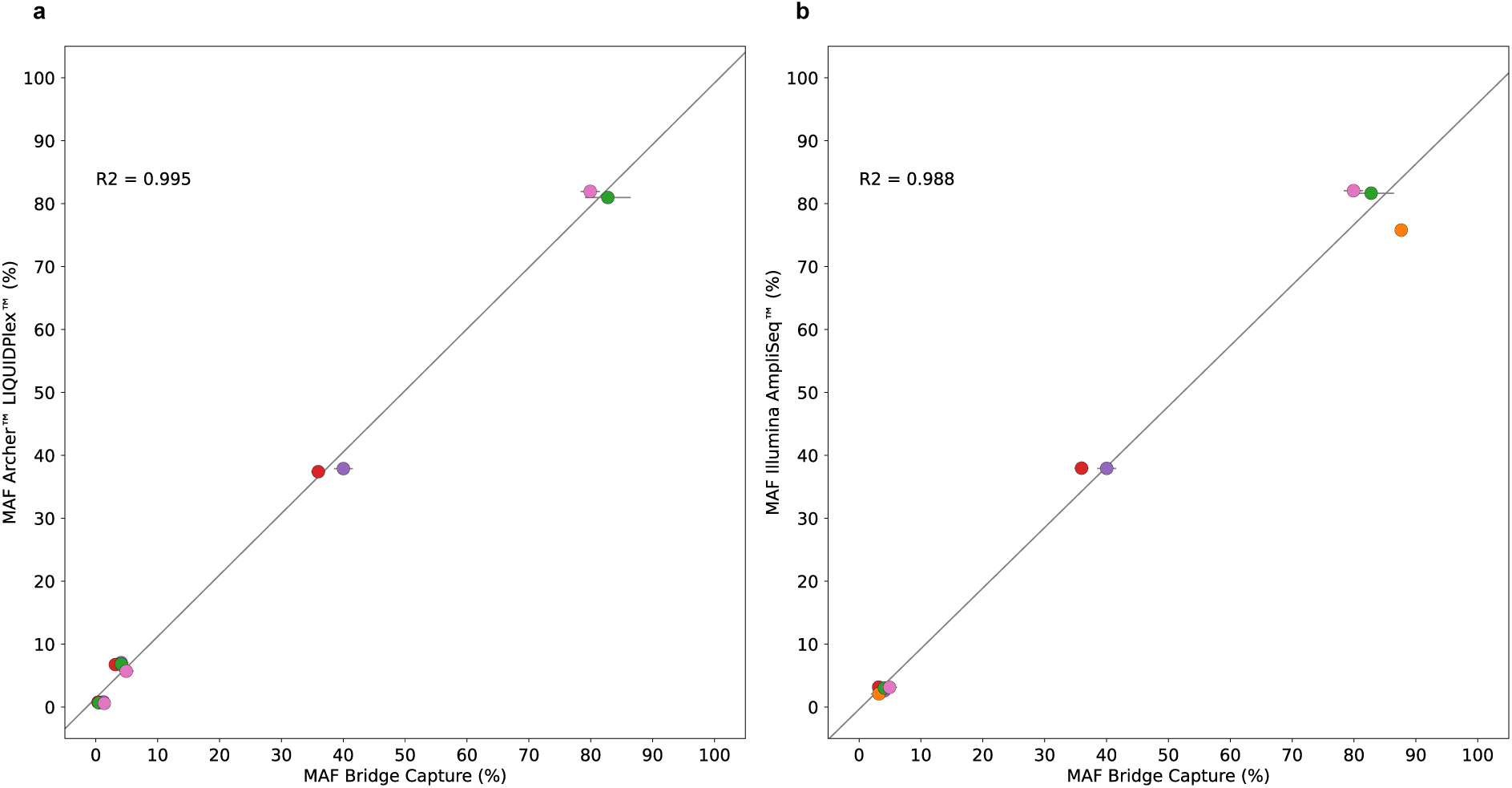
Concordance between MAFs detected by Bridge Capture and commercially available technologies. (**a**) Concordance between MAFs detected by Archer™ LIQUIDPlex™ assay and corresponding MAFs detected by Bridge Capture. (**b**) Concordance between MAFs detected by AmpliSeq™ Cancer HotSpot Panel v2 for Illumina® and corresponding MAFs detected by Bridge Capture. The mutations are indicated as follows: APC p.Q1406* (blue), KRAS p.G13D (purple), PIK3CA p.E545K (red), APC p.E1309Dfs*4 (orange), KRAS p.G12D (green), TP53 p.Y126D (pink).

All technologies consistently detected SNVs > 2 % MAF (**Table 1**). The lowest MAF identified by Bridge Capture, LIQUIDPlex and AmpliSeq were 0.08 %, 0.58 % and 2.0 %, respectively. The only deletion (*APC* p.E1309Dfs*4) was detected by both Bridge Capture and AmpliSeq at MAF above 2 %. All technologies exhibited high linear correlation between the sample dilution factor and MAF, implying consistent linear performance across wide range of MAFs. Average R^2^ of the detected variants was 0.961, 0.999, and 0.995 for Bridge Capture, Ampliseq, and LIQUIDPlex, respectively.

**Table 1.**
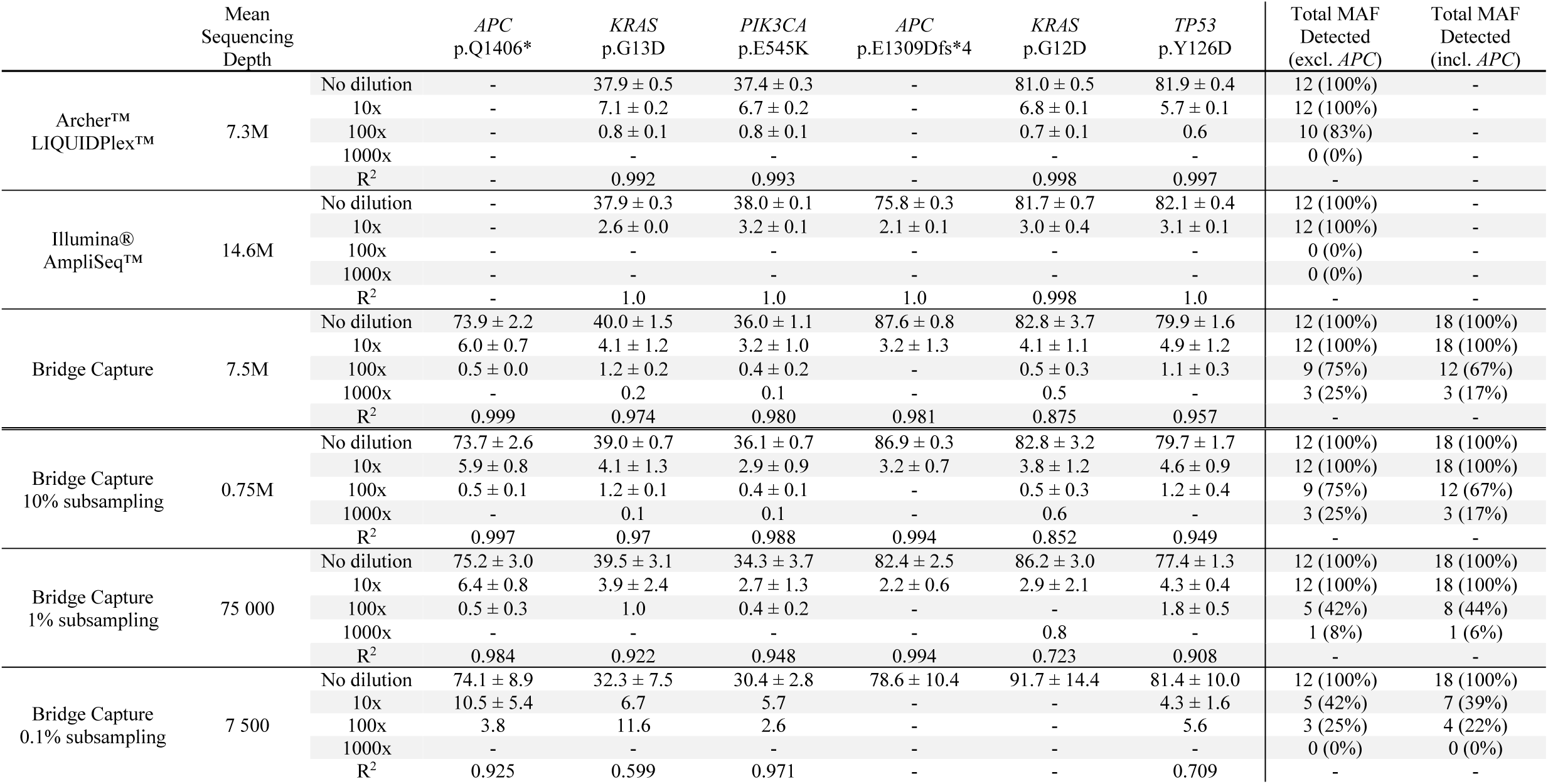
Comparison of MAFs detected by Archer™ LIQUIDPlex™, AmpliSeq™ Cancer HotSpot Panel v2 for Illumina®, and Bridge Capture; including subsampling analysis of Bridge Capture. The MAFs are shown for respective methods across different sample dilutions (No dilution, 10×, 100×, 1000×) with corresponding R^2^ value for each specified gene and its variant. The MAFs are presented as mean ± standard deviation from three replicates, although some variants were not detected in all replicates. For Bridge Capture, the raw reads obtained by sequencing (7.5M reads) were subsampled by 10 % (0.75M reads), 1 % (75,000 reads) and 0.1 % (7500 reads), highlighting the effect of sequencing depth on MAF detection. Table summarizes the total MAF detected excluding APC and including APC for given sample dilution. Total MAF detected including APC are given only for Bridge Capture since the other methods didn’t cover all APC variants in their panel.

|  | Mean Sequencing Depth |  | *APC*<br>p.Q1406* | *KRAS*<br>p.G13D | *PIK3CA*<br>p.E545K | *APC*<br>p.E1309Dfs*4 | *KRAS*<br>p.G12D | *TP53*<br>p.Y126D | Total MAF Detected<br>(excl. *APC*) | Total MAF Detected<br>(incl. *APC*) |
| --- | --- | --- | --- | --- | --- | --- | --- | --- | --- | --- |
| Archer™<br>LIQUIDPlex™ | 7.3M | No dilution | - | 37.9 ± 0.5 | 37.4 ± 0.3 | - | 81.0 ± 0.5 | 81.9 ± 0.4 | 12 (100%) | - |
|  |  | 10x | - | 7.1 ± 0.2 | 6.7 ± 0.2 | - | 6.8 ± 0.1 | 5.7 ± 0.1 | 12 (100%) | - |
|  |  | 100x | - | 0.8 ± 0.1 | 0.8 ± 0.1 | - | 0.7 ± 0.1 | 0.6 | 10 (83%) | - |
|  |  | 1000x | - | - | - | - | - | - | 0 (0%) | - |
|  |  | R<sup>2</sup> | - | 0.992 | 0.993 | - | 0.998 | 0.997 | - | - |
| Illumina®<br>AmpliSeq™ | 14.6M | No dilution | - | 37.9 ± 0.3 | 38.0 ± 0.1 | 75.8 ± 0.3 | 81.7 ± 0.7 | 82.1 ± 0.4 | 12 (100%) | - |
|  |  | 10x | - | 2.6 ± 0.0 | 3.2 ± 0.1 | 2.1 ± 0.1 | 3.0 ± 0.4 | 3.1 ± 0.1 | 12 (100%) | - |
|  |  | 100x | - | - | - | - | - | - | 0 (0%) | - |
|  |  | 1000x | - | - | - | - | - | - | 0 (0%) | - |
|  |  | R<sup>2</sup> | - | 1.0 | 1.0 | 1.0 | 0.998 | 1.0 | - | - |
| Bridge Capture | 7.5M | No dilution | 73.9 ± 2.2 | 40.0 ± 1.5 | 36.0 ± 1.1 | 87.6 ± 0.8 | 82.8 ± 3.7 | 79.9 ± 1.6 | 12 (100%) | 18 (100%) |
|  |  | 10x | 6.0 ± 0.7 | 4.1 ± 1.2 | 3.2 ± 1.0 | 3.2 ± 1.3 | 4.1 ± 1.1 | 4.9 ± 1.2 | 12 (100%) | 18 (100%) |
|  |  | 100x | 0.5 ± 0.0 | 1.2 ± 0.2 | 0.4 ± 0.2 | - | 0.5 ± 0.3 | 1.1 ± 0.3 | 9 (75%) | 12 (67%) |
|  |  | 1000x | - | 0.2 | 0.1 | - | 0.5 | - | 3 (25%) | 3 (17%) |
|  |  | R<sup>2</sup> | 0.999 | 0.974 | 0.980 | 0.981 | 0.875 | 0.957 | - | - |
| Bridge Capture<br>10% subsampling | 0.75M | No dilution | 73.7 ± 2.6 | 39.0 ± 0.7 | 36.1 ± 0.7 | 86.9 ± 0.3 | 82.8 ± 3.2 | 79.7 ± 1.7 | 12 (100%) | 18 (100%) |
|  |  | 10x | 5.9 ± 0.8 | 4.1 ± 1.3 | 2.9 ± 0.9 | 3.2 ± 0.7 | 3.8 ± 1.2 | 4.6 ± 0.9 | 12 (100%) | 18 (100%) |
|  |  | 100x | 0.5 ± 0.1 | 1.2 ± 0.1 | 0.4 ± 0.1 | - | 0.5 ± 0.3 | 1.2 ± 0.4 | 9 (75%) | 12 (67%) |
|  |  | 1000x | - | 0.1 | 0.1 | - | 0.6 | - | 3 (25%) | 3 (17%) |
|  |  | R<sup>2</sup> | 0.997 | 0.97 | 0.988 | 0.994 | 0.852 | 0.949 | - | - |
| Bridge Capture<br>1% subsampling | 75 000 | No dilution | 75.2 ± 3.0 | 39.5 ± 3.1 | 34.3 ± 3.7 | 82.4 ± 2.5 | 86.2 ± 3.0 | 77.4 ± 1.3 | 12 (100%) | 18 (100%) |
|  |  | 10x | 6.4 ± 0.8 | 3.9 ± 2.4 | 2.7 ± 1.3 | 2.2 ± 0.6 | 2.9 ± 2.1 | 4.3 ± 0.4 | 12 (100%) | 18 (100%) |
|  |  | 100x | 0.5 ± 0.3 | 1.0 | 0.4 ± 0.2 | - | - | 1.8 ± 0.5 | 5 (42%) | 8 (44%) |
|  |  | 1000x | - | - | - | - | 0.8 | - | 1 (8%) | 1 (6%) |
|  |  | R<sup>2</sup> | 0.984 | 0.922 | 0.948 | 0.994 | 0.723 | 0.908 | - | - |
| Bridge Capture<br>0.1% subsampling | 7 500 | No dilution | 74.1 ± 8.9 | 32.3 ± 7.5 | 30.4 ± 2.8 | 78.6 ± 10.4 | 91.7 ± 14.4 | 81.4 ± 10.0 | 12 (100%) | 18 (100%) |
|  |  | 10x | 10.5 ± 5.4 | 6.7 | 5.7 | - | - | 4.3 ± 1.6 | 5 (42%) | 7 (39%) |
|  |  | 100x | 3.8 | 11.6 | 2.6 | - | - | 5.6 | 3 (25%) | 4 (22%) |
|  |  | 1000x | - | - | - | - | - | - | 0 (0%) | 0 (0%) |
|  |  | R<sup>2</sup> | 0.925 | 0.599 | 0.971 | - | - | 0.709 | - | - |

To determine the sequencing depth required by Bridge Capture for detecting low MAFs, the raw sequencing data (7.5M reads) was subsampled (**Table 1**). At 10 % subsampling, corresponding to approximately 750,000 reads, Bridge Capture could detect MAFs below 0.1 % for *KRAS* p.G13D, *PIK3CA* p.E545K and *KRAS* p.G12D and results were nearly identical with the non-subsampled data. At 1 % subsampling, corresponding to approximately 75,000 reads, Bridge Capture could detect MAFs down to 1 % for all mutations except *APC* p.E1309Dfs*4.

### Inter-lab reproducibility of Bridge Capture

The portability and reproducibility of Bridge Capture was assessed by comparing the results of an assay performed by an independent diagnostic service provider in the UK (site A) to the results from Genomill laboratory (site B) (**Fig. 3a**). The MAFs detected between sites A and B were strongly correlated (R^2^ = 0.979).

**Figure 3.**
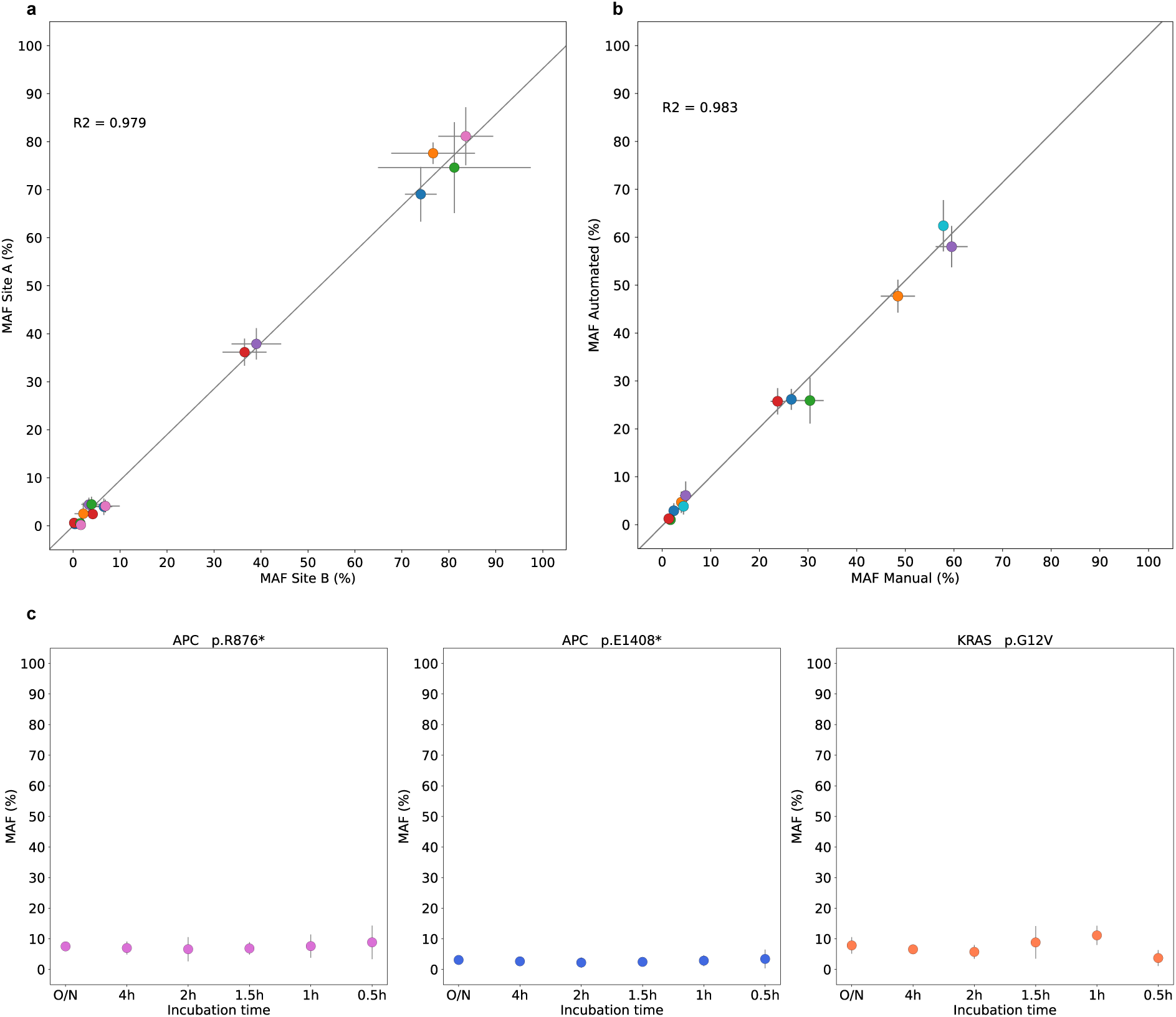
(**a**) The inter-lab reproducibility testing of Bridge Capture between an independent diagnostic service provider laboratory site in UK (site A) and Genomill laboratory, Turku, Finland (site B), shows a high correlation (R^2^ = 0.979). The mutations are indicated as follows: APC p.Q1406* (blue), KRAS p.G13D (purple), PIK3CA p.E545K (red), APC p.E1309Dfs*4 (orange), KRAS p.G12D (green), TP53 p.Y126D (pink). (**b**) The reproducibility of Bridge Capture between automated and manual workflow is high, as shown by the strong linear correlation between the workflows (R^2^ = 0.983, r = 0.992). The mutations are indicated as follows: APC p.R876* (purple), APC p.E1408* (red), CTNNB1 p.S45P (blue), KRAS p.G12V (cyan), KRAS p.G13D (orange), TP53 p.R248W (green) (**c**) Hybridization time (MAF based on 5 replicates) had a significant effect on *KRAS* p.G12V (one-way ANOVA, p < 0.05) and did not have a significant effect on the other mutations. The points are represented as the mean of the replicas ± standard deviation.

### Integration of automated Bridge Capture workflow

The implementation of automated Bridge Capture workflow was evaluated by comparison to manual workflow using two contrived CRC patient samples, tested in three replicates with both workflows. The MAFs obtained from the automated and manual workflow were highly correlated (R^2^ = 0.983, r = 0.992) (**Fig. 3b**).

The hands-on time required for processing 15 samples was recorded for both workflows. The basic set up of the automated workflow took 4 minutes and 20 seconds (positioning of the labware, addition of ice and reagents for the first workflow step, final robot clean-up) and subsequent removal and addition of reagents took 6 minutes and 18 seconds. This additional step was obligatory, as the pipetting robot lacks cooling module, and reagents must be added separately during each step of workflow. However, with more advanced pipetting robot, this step would not be required. Therefore, the total hands-on time for automated workflow was 10 minutes and 38 seconds. For manual workflow, the hands-on time was 35:15 minutes excluding the bead-purification of the ready-made libraries before the sequencing.

### The effect of hybridization time on detection limit of Bridge Capture

The effect of hybridization time on Bridge Capture performance was tested by detecting MAFs using six different incubation times (O/N, 4 h, 2 h, 1.5 h, 1 h, 0.5 h) in five replicates (**Fig. 3c**). Mutations of interest were successfully identified in all replicates, excluding one replicate in the 0.5 h incubation time point, where the *APC* E1408* was not detected. The standard deviation of the MAFs detected between the five replicates was the lowest for replicates incubated O/N and 4 h. The standard deviation between replicates was increasing with shorter incubation time and its peak could be observed at 0.5 h time point. Only the detection of *KRAS* p.G12V was significantly affected by changes in the incubation time (one-way ANOVA test, p < 0.05).

### Panel scalability

To assess the scalability of the Bridge Capture method, we increased the panel size by more than 300% from 282 to 887 probes. The effect of the panel size increase was explored by evaluating the change in probe signal intensity and the performance of the matching probes between the two panels. Signal intensity was established for each probe from ten replicates of fragmented gDNA (**Fig. 4a** and **Fig. 4b**) for panels of 282 and 887 probes, by dividing the read count per probe by the total read count of each replicate. Probe signal intensity was varying between the probes, with median and mean intensity of 0.324 and 0.355, compared to theoretical intensity of 0.355 assuming a uniform performance for the 282-probe panel. In comparison, for the 887-probe panel, the median and mean intensity was 0.087 and 0.113, compared to the theoretical intensity of 0.087. Signal intensity per probe between replicates was notably consistent, with a mean standard deviation of 0.0267 for the 282-probe panel (p < 0.001) and 0.0091 for the 887-probe panel (p < 0.001).

**Figure 4.**
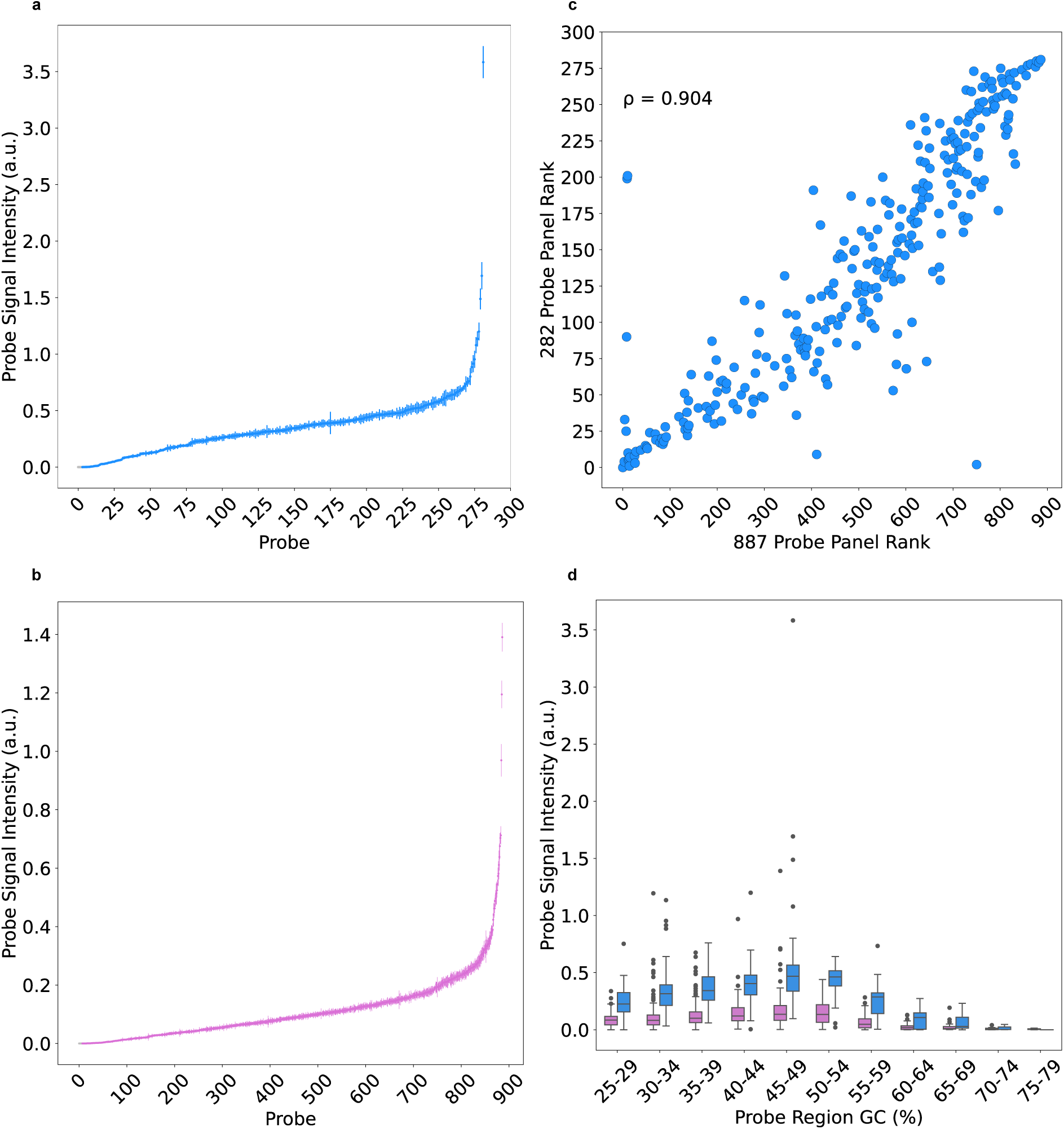
Probe signal intensity of (**a**) 282-probe panel and (**b**) 887-probe panel. Mean probe signal intensity of ten replicates (y-axis) was calculated by dividing the read count per probe by the total read count of the replicate. Standard deviation between the replicates is displayed as error bar. **(c)** Spearman’s rank correlation (ρ = 0.904) depicting the performance relationship of the probes shared between 282- and 887-probe panel. (**d**) The relationship between probe signal intensity and GC-content of the probe target region (300 nt around the probe binding site) of 282-probe panel (blue) and 887-probe panel (pink).

The probes present in both panels performed highly similarly as indicated by Spearman’s rank correlation (ϱ = 0.904), demonstrating that panel size expansion had negligible impact on performance and cross-reactivity of individual probes (**Fig. 4c)**.

A relationship between probe signal intensity and the respective GC-content of the target region (defined as 300 nt around the probe binding site) was observed (**Fig. 4d**). For both panels, high GC-content affected probe performance evidenced by a dip in signal intensity starting from 55 – 59 % with a noticeable decline from 60 % onwards. Similarly, but to a lesser degree, low GC-content (25 – 29 %) had a negative impact on probe performance as well.

The 282-probe panel covers 64 cancer types and associated driver genes obtained from intOgen-framework(22) and their respective mutations of 1 and 2 significance which are based on COSMIC database, CMC v.99(23). A majority of cancer type mutations are covered by the 282-probe panel (**Supplementary Figure 1**). Expanded 887-probe panel covers nearly 71 driver genes with a majority of the cancer types (65/71) covered by at least 90 %. This panel includes probes for most common cancer types such as colorectal adenocarcinoma, breast carcinoma, non-small cell lung cancer, prostate adenocarcinoma, and ovarian cancer.

## Discussion

We demonstrate that Bridge Capture surpasses the performance of commercially available leading technologies for cancer diagnostics such as LIQUIDPlex and AmpliSeq. Furthermore, Bridge Capture addresses the challenges of hybridization-, amplicon-and MIP-based technologies and offers a simple, cost-efficient, single-day, automatable workflow with high performance. For example, Bridge Capture identified certain mutations that were below the cutoff limit of LIQUIDPlex (> 0.5 %) and AmpliSeq (> 1 %). We also demonstrated that Bridge Capture is easily adoptable in a new laboratory setting, shown by the high correlation between two independent laboratories. This was achieved despite the use of different sequencing platforms, over 9-fold difference in sequencing depth, and operators having varying levels of experience.

Various library preparation methods are available for NGS-based liquid biopsy analysis, each with their advantages and limitations. For instance, amplicon-based technologies such as AmpliSeq and TruSeq® Amplicon are simple and rapid in terms of their workflow but provide limited scalability to a larger number of targets (11–13). Hybrid capture methods such as AVENIO and FoundationACT provide scalability to a very large number of target genes but are slow, cumbersome and expensive (16,17,24,25). MIP-based workflows are rapid and simple but suffer from high probe synthesis costs as well as poor uniformity (19,26). The results presented in this research provide initial evidence that Bridge Capture based sequencing has the potential to combine the advantages observed in amplicon- and hybrid capture-based methods.

The sequencing depth required by Bridge Capture is low, with approximately 750,000 reads sufficient for detecting mutations of 0.1 % MAF with a panel of 282 probes. The low sequencing depth requirement permits pooling dozens of samples even on benchtop devices such as Illumina MiniSeq, and more than thousands of samples on production-scale sequencers such as NovaSeq X. This could potentially be leveraged in small and medium scale hospitals and laboratories where the weekly or monthly sample volume would not be sufficient for filling up runs on production scale sequencers or enabling higher throughput in larger hospitals and centralized laboratories.

We demonstrate panel expansion from 282 to 887 probes. Despite the panel coverage increasing by more than 300 %, the panel uniformity remains unchanged, implying that the Bridge Capture panels can be scaled without any theoretical or practical upper limit. However, similar to other available methods, the performance between probes can be variable, with certain probes over- or underperforming, which can be mitigated by probe design. We observed that the probe performance is related to the high or low GC-content of the probe target region. This performance effect could result from the indexing PCR(27–30), since PCR has been shown to deplete loci with GC-content of more than 65 % (29,30). Another potential source for the probe performance imbalance could be the sequencing by synthesis on Illumina platforms, which is also negatively impacted by high GC-content (28,29,31). Outliers above the average performance could partly be explained by the binding site overrepresentation by pseudogenes, for instance the probe targeting NF1 gene, which is a known homologous pseudogene located through the human genome (32,33).

We expect to further increase the sensitivity of Bridge Capture by refining our probe design and optimizing the reaction process to exhaustively target all DNA molecules in a sample, as well as to extend the validation to cover copy number variations and fusions. Bridge Capture provides a scalable but highly targeted approach to molecular diagnostics, as with the 887-probe panel and low sequencing depth requirements we are already able to address the most common cancer types such as colorectal adenocarcinoma, breast carcinoma, non-small cell lung cancer, prostate adenocarcinoma, and ovarian cancer.

In conclusion, Bridge Capture is an easy-to-use, cost-efficient, scalable, streamlined and platform-agnostic NGS library preparation method that permits combining the speed and simplicity of the amplicon- and MIP-based methods while maintaining the performance with the scalability and sensitivity of the hybrid capture methods. Hence, Bridge Capture is a novel cancer diagnostics tool that significantly improves the detection of mutations in liquid biopsies.

## Abbreviations

ANOVA: Analysis of variance
cfDNA: Cell-free DNA
CRC: Colorectal cancer
ctDNA: Circulating tumor DNA
dsDNA: Double-stranded DNA
gDNA: Genomic DNA
MAF: Mutant allele frequency
MIP: Molecular inversion probe
NGS: Next-generation sequencing
O/N: Overnight
PCR: Polymerase chain reaction
RCA: Rolling circle amplification
SNV: Single-nucleotide variant
ssDNA: Single-stranded DNA
UMI: Unique molecular identifier
WGS: Whole genome sequencing

## Data availability

The data underlying this article will be shared on reasonable request to the corresponding author.

## Acknowledgements

The authors would like to express their gratitude to Voima Ventures, Avohoidon Tutkimussäätiö and Almaral for support and funding.

