## Supplemental tables and figures for "Bridge Capture permits cost-efficient, rapid and sensitive molecular precision diagnostics"

**Supplementary Table 1.** Composition of 282-probe panel. Table represent the list of genes included in the panel, alongside the corresponding number of probes.

**84-gene panel consisting of 282 probes.**

| Gene | Probes | Gene | Probes | Gene | Probes | Gene | Probes |
|---|---|---|---|---|---|---|---|
| ABL1 | 1 | CTNNB1 | 2 | KMT2D | 5 | PTPN11 | 1 |
| ALK | 4 | CUL3 | 2 | KRAS | 3 | RAC1 | 1 |
| APC | 39 | DICER1 | 1 | MAP2K1 | 1 | RAF1 | 1 |
| AR | 1 | DNMT3A | 7 | MET | 1 | RB1 | 12 |
| ASXL1 | 6 | EGFR | 4 | MLH1 | 6 | RBM10 | 1 |
| ATM | 11 | EP300 | 1 | MSH2 | 10 | RUNX1 | 1 |
| ATR | 1 | ERBB2 | 1 | MSH6 | 2 | SETD2 | 1 |
| B2M | 1 | ERBB3 | 2 | MTOR | 1 | SF3B1 | 3 |
| BAP1 | 1 | EZH2 | 1 | MUTYH | 1 | SMAD2 | 1 |
| BCL2L2 | 1 | FANCA | 1 | NBN | 1 | SMAD4 | 9 |
| BRAF | 2 | FBXW7 | 5 | NF1 | 18 | SPEN | 1 |
| BRCA1 | 6 | FGFR2 | 2 | NF2 | 2 | SPOP | 2 |
| BRCA2 | 5 | FGFR3 | 1 | NFE2L2 | 2 | STAG2 | 1 |
| CBL | 1 | FLT3 | 2 | NFKBIA | 1 | STK11 | 2 |
| CDC73 | 1 | GNAQ | 1 | NOTCH1 | 4 | TERT | 1 |
| CDH1 | 3 | GNAS | 2 | NRAS | 2 | TP53 | 16 |
| CDK12 | 1 | HRAS | 1 | PALB2 | 3 | TSC1 | 2 |
| CDKN1B | 1 | IDH1 | 1 | PDGFRA | 1 | U2AF1 | 2 |
| CDKN2A | 3 | JAK2 | 1 | PIK3CA | 4 | VHL | 4 |
| CHEK2 | 5 | KEAP1 | 1 | PTCH1 | 3 | WT1 | 1 |
| CREBBP | 2 | KIT | 2 | PTEN | 10 | XPO1 | 1 |

**Supplementary Table 2.** Composition of 887-probe panel. Table represent the list of genes included in the panel, alongside the corresponding number of probes.

| **123-gene panel consisting of 887 probes.** | | | | | | | |
|---|---|---|---|---|---|---|---|
| Gene | Probes | Gene | Probes | Gene | Probes | Gene | Probes |
| ABL1 | 2 | CTNNB1 | 2 | KEAP1 | 1 | PPP2R1A | 1 |
| AKT1 | 1 | CUL3 | 2 | KIT | 2 | PTCH1 | 3 |
| ALK | 4 | CYLD | 4 | KLF4 | 1 | PTEN | 19 |
| APC | 81 | DICER1 | 12 | KMT2D | 6 | PTPN11 | 3 |
| AR | 3 | DNMT3A | 9 | KRAS | 3 | RAC1 | 1 |
| ARID1A | 1 | EGFR | 9 | LZTR1 | 1 | RAF1 | 1 |
| ARID1B | 1 | EIF3E | 1 | MAP2K1 | 2 | RB1 | 36 |
| ASXL1 | 6 | EP300 | 9 | MAP3K1 | 1 | RBM10 | 1 |
| ATM | 83 | ERBB2 | 6 | MAPK1 | 1 | RET | 1 |
| ATR | 1 | ERBB3 | 4 | MAX | 4 | RHOA | 1 |
| B2M | 1 | ESR1 | 1 | MEN1 | 8 | RNF43 | 2 |
| BAP1 | 17 | EZH2 | 1 | MET | 1 | RUNX1 | 8 |
| BCL2L2 | 1 | FANCA | 1 | MLH1 | 8 | SETD2 | 3 |
| BCL9 | 1 | FBXW7 | 5 | MSH2 | 11 | SF3B1 | 4 |
| BCOR | 1 | FGFR1 | 2 | MSH6 | 3 | SMAD2 | 1 |
| BMPR1A | 7 | FGFR2 | 4 | MTOR | 4 | SMAD4 | 18 |
| BRAF | 3 | FGFR3 | 3 | MUTYH | 1 | SMARCA4 | 16 |
| BRCA1 | 47 | FH | 1 | MYC | 1 | SPEN | 1 |
| BRCA2 | 70 | FLCN | 5 | NBN | 1 | SPOP | 2 |
| BTK | 1 | FLT3 | 2 | NF1 | 100 | STAG2 | 1 |
| CASP8 | 3 | GATA1 | 1 | NF2 | 14 | STK11 | 14 |
| CBL | 1 | GATA3 | 1 | NFE2L2 | 2 | TERT | 1 |
| CDC73 | 1 | GNA11 | 1 | NFKBIA | 1 | TGFBR2 | 1 |
| CDH1 | 19 | GNAQ | 2 | NOTCH1 | 5 | TP53 | 19 |
| CDK12 | 1 | GNAS | 2 | NOTCH2 | 1 | TSC1 | 2 |
| CDK4 | 1 | HRAS | 2 | NRAS | 2 | TSC2 | 1 |
| CDKN1B | 2 | IDH1 | 1 | PALB2 | 3 | U2AF1 | 2 |
| CDKN2A | 8 | IDH2 | 2 | PDGFRA | 1 | VHL | 9 |
| CHEK2 | 5 | IRS4 | 1 | PIK3CA | 13 | WT1 | 1 |
| CREBBP | 23 | JAK2 | 1 | PIK3R1 | 4 | XPO1 | 1 |
| CTCF | 1 | KDM6A | 1 | PMS2 | 12 | | |

**Supplementary Table 3.** List of the contrived CRC samples analysed by Bridge Capture, Archer™ LIQUIDPlex™ and AmpliSeq™ Cancer HotSpot Panel v2 for Illumina® to evaluate the concordance between technologies and establish the detection limits of the technologies. Table describes composition of each contrived sample and displays the representation of original patient MAFs after the dilution with genomic DNA.

| **Experimental design** | | | | |
|---|---|---|---|---|
| **Sample number** | **Patient** | **Patient copies** | **Healthy genomic DNA copies** | **Expected mutations** |
| 1<br>2<br>3 | Patient 1 | 100 000 | - | 100% of original MAF of APC p.Q1406*, KRAS p.G13D, PIK3CA p.E545K |
| 4<br>5<br>6 | Patient 1 | 10 000 | 90 000 | 10% of original MAF of APC p.Q1406*, KRAS p.G13D, PIK3CA p.E545K |
| 7<br>8<br>9 | Patient 1 | 1 000 | 100 000 | 1% of original MAF of APC p.Q1406*, KRAS p.G13D, PIK3CA p.E545K |
| 10<br>11<br>12 | Patient 1 | 100 | 100 000 | 0.1% of original MAF of APC p.Q1406*, p.G13D, PIK3CA p.E545K |
| 13<br>14<br>15 | Patient 2 | 100 000 | - | 100% of original MAF of APC p.E1309Dfs*4, KRAS p.G12D, TP53 p.Y126D |
| 16<br>17<br>18 | Patient 2 | 10 000 | 90 000 | 10% of original MAF of APC p.E1309Dfs*4, KRAS p.G12D, TP53 p.Y126D |
| 19<br>20<br>21 | Patient 2 | 1 000 | 100 000 | 1% of original MAF of APC p.E1309Dfs*4, KRAS p.G12D, TP53 p.Y126D |
| 22<br>23<br>24 | Patient 2 | 100 | 100 000 | 0.1% of original MAF of APC p.E1309Dfs*4, KRAS p.G12D, TP53 p.Y126D |

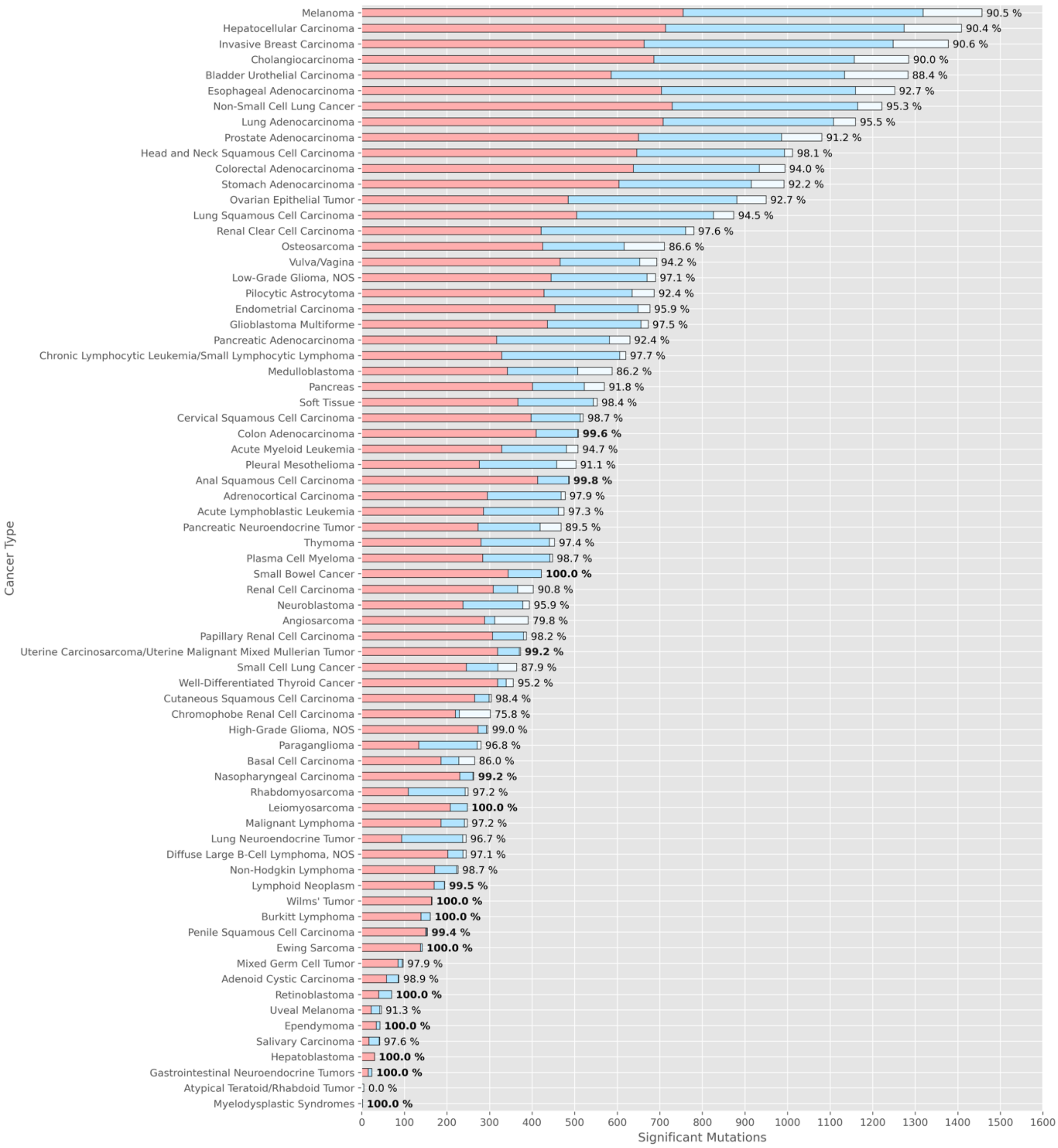


**Supplementary Figure 1.** The expansion of 282-probe panel to 887-probe panel. The bars indicates the number of significant mutations associated with driver genes in 282-probe panel (red) and 887-probe panel (blue). White bars represent non-targeted significant mutations of the driver genes not included in either of the panels.
The percentage is a ratio of number of significant mutations targeted by the 887-probe panel and total amount of significant mutations for that specific cancer type. Cancer types and their associated driver genes are obtained from intOgen-framework (https://www.intogen.org) v.2023.05.31. Significant mutations of the driver genes are mutations of significance 1 or 2 in COSMIC database, CMC v.99 (https://cancer.sanger.ac.uk/cosmic).
